# The Prevalence of the Female Athlete Triad in Female Service Members

**DOI:** 10.1101/2023.04.06.23288270

**Authors:** Cameron S. Mattison, John Wiese

**Affiliations:** Medical College of Wisconsin

## Abstract

Osteoporosis, amenorrhea, and low energy with or without disordered eating (the female athlete triad) are frequent clinical outcomes associated with female athletes in constant low energy availability (LEA). The rigorous training demands of the Army and the strict weight limits suggest that female service members may be susceptible to states of LEA, where energy expenditure exceeds dietary energy intake.

To understand the prevalence of the female athlete triad among female service members and how restrictive weight requirements influence these symptoms, we compared survey responses measuring symptoms of the female athlete triad between female active-duty service members and female veterans.

The results indicated that female veterans had significantly higher female athlete triad and disordered eating scores. Therefore, female veterans are more likely to demonstrate female athlete triad symptoms than active-duty service members. Specifically, female veterans are more likely to demonstrate symptoms of low energy with or without disordered eating.

Female veterans’ unique stressors of no longer being in the military (reintegration into society and losing support system within the military) could be why they are more likely to present with female athlete triad symptoms than active-duty female service members.

## Introduction

When energy availability is consistently imbalanced, where energy expenditure (energy required for exercise or basic metabolic function outweighs dietary energy intake, basic metabolic functions are impaired.^1^ This state of being in an energy intake deficit is a period of low energy availability (LEA). Since high energy expenditure and weight-sensitive sports seem to contribute to the prevalence of LEA among athletes, the high training demands of the Army and stringent weight requirements suggest that U.S. Army soldiers may be predisposed to states of LEA.^2^

A common clinical result linked to persistent LEA is osteoporosis, amenorrhea, and disorder eating habits.^3^ These clinical symptoms presented in female athletes in continuous states of LEA have been termed the female athlete triad. The current weight standard for female soldiers—the primary goal that every soldier should meet—is stricter than for male soldiers.^4^ These weight restrictions and the physical demands of the Army may predispose female soldiers to the female athlete triad, which includes low energy availability (EA) with or without disordered eating (DE), menstrual disruptions (MD), and poor bone mineral density (BMD).

We expect that the greater pressure that female active-duty service members have to adhere to restrictive weight requirements will reveal that female active-duty service members experience more components of the female athlete triad, including EA with or without DE, MD, and low BMD compared to female veterans. Research on the prevalence of the female athlete triad among female military service members is scarce. Therefore, studies investigating the prevalence of the triad among female service members can provide physicians valuable insight into whether this is a population of concern for female athlete triad symptoms.

## Method

This research project is a 4×4 between subjects design. We collected data through an online survey that was distributed to 4,000 female Army service members (total combination of veterans and active-duty Army personnel) through the West Point Women’s Facebook Group Page. This Facebook group page requires that every member provides identification that they are either graduates or cadets at West Point, an institution where every student serves in the United States Army, before being accepted as a member of the group.

Every participant conducted a survey that measured components of the female athlete triad, including EA with or without disordered eating DE, MD, and low BMD. The survey questions were based on the Female Athlete Screening Tool (FAST) and the Low Energy Availability in Female Questionnaire (LEAF-Q). Researchers have devised and validated the LEAF-Q as a screening tool for female athletes.^5^ This survey can identify female athletes “at risk” of the physiological signs linked to LEA with a 78% sensitivity and 90% specificity. While it can be utilized alone, it is advised that it be used in conjunction with a validated DE screening instrument, such as the Female Athlete Screening Tool (FAST).^6^

We excluded participants who were currently pregnant, post-menopause, and currently taking birth control/medication that prevented them from having regular periods. A total of 64 participants (51 active-duty female service members and 13 female veterans).

To analyze the data, we conducted a four by four, between subjects ANOVA test through SPSS. The ANOVA test analyzed the differences of survey responses between military status (female active-duty service members and female veterans) regarding a female athlete triad score, and subcomponent scores of the female athlete triad (disordered eating, amenorrhea, and osteoporosis).

## Results

The ANOVA tests revealed that the responses between female active duty service members and female veterans regarding disordered eating F(1, 64) = 6.78, *p* = 0.01, and the overall female athlete triad score showed significant differences F(1, 64) = 7.06, *p* = 0.01. The female veterans demonstrated higher disordered eating behaviors, (*M*=174.192, *SD*=23.526), compared to female active-duty service members (*M*=141.510, *SD*=43.489). The female veterans also had a higher female athlete triad score (*M*=212.023, *SD*=25.122) compared to female service members (*M*=177.822, *SD*=44.448). See Table 1 for all values.

**Table 1.** Differences in Responses for Female Athlete Triad and Sub-Components (Disordered Eating, Amenorrhea, and Osteoporosis)

|  | Military Status | Mean | Standard<br>Deviation |
| --- | --- | --- | --- |
| Female Athlete<br>Triad Score* | Veteran | 212.02 | 25.12 |
|  | Active Duty | 177.82 | 44.45 |
| Disordered Eating<br>Score* | Veteran | 174.19 | 23.52 |
|  | Active Duty | 141.51 | 43.49 |
| Amenorrhea Score | Veteran | 13.18 | 4.32 |
|  | Active Duty | 12.94 | 2.87 |
| Osteoporosis Score | Veteran | 24.65 | 6.81 |
|  | Active Duty | 23.37 | 5.81 |

\* Indicates significance

To better understand our results, we conducted more statistical tests regarding specific questions in the survey. We conducted another one-way ANOVA test between female active-duty service members and female veterans regarding their responses to, “I believe most of my eating habits have been influenced by the diet culture in the Army” (Q1). This test revealed that there was a significant difference in responses regarding Q1, F(1, 64) = 4.55, *p* = 0.04. Specifically, female veterans seemed to agree with the statement in Q1 (*M*=8.92, *SD*=1.26) more than female active-duty service members (M=6.86, *SD*=3.41). We also tested for descriptive statics regarding responses from female active-duty service members with the questions, “I am concerned about meeting Army height and weight requirements” (Q2) and “I have done things to keep my weight down that I believe are unhealthy” (Q3). These descriptive statistics show that female active-duty service members had a *M*=*7*.*38, SD*=*3*.*36* to Q2 and a *M*=*7*.*67, SD=3*.*23* to Q3.

## Discussion

The findings contradict our hypothesis that, when compared to female veterans, female active-duty military members are more likely to suffer from EA with or without DE, MD, and low BMD. Our findings specifically showed that female veterans had a higher overall female athlete triad score and considerably higher disordered eating score than female active-duty military members.

Our female veterans reported a higher level of agreement with the statement that the Army’s diet culture influenced their eating habits, despite no longer being subjected to the Army’s stringent weight standards and high energy demands. Therefore, they indicated that their military experiences had a long-lasting effect on their eating habits, which may explain why the female veterans showed a high disordered eating score.

Research regarding different types of traumas associated with the prevalence of eating disorders in female veterans also offers insight into our findings. According to this study, disordered eating is associated with military-related trauma, and the general stress brought on by these traumas is connected with an increase in disordered eating symptoms.^7^ While female veterans and active-duty service members may have experienced similar military traumas that contributed to their disordered eating habits, female veterans may have added stressors related to their veteran status. These unique stressors include reintegration into civilian society and losing any support system they had while in the military. These added stressors experienced by female veterans may cause their higher disordered eating score, leading to a higher female athlete triad score than active-duty female service members.

Even though the female veterans scored higher on the female athlete triad and disordered eating scales, several active-duty female service members’ answers raised concerns. In particular, female active-duty service members mostly agreed with the statement that they were concerned about fulfilling the Army’s height and weight limits and that they had engaged in actions they thought to be unhealthy to maintain their weight (see Figures 2 and 3). These responses suggest that female active-duty service members may still be in stages of LEA, but their symptoms corresponding to the female athlete triad are just not as prevalent as their female veteran counterparts.

There should be a further investigation that compares female active duty and female veterans to other female athletes. This further investigation will provide more insight into the severity of the female athlete triad symptoms in the female service member population. A more extensive project that uses lab measures to evaluate bone mineral density will also provide a more accurate understanding of the bone mineral density in these populations. Our limited means to contact female veterans also hindered this project’s ability to provide a generalized understanding of the prevalence of the female athlete triad in this population. Despite having limited resources confining our study to surveys rather than labs, and limited responses from female veterans, this study serves as a starting point in investigating the female athlete triad among female service members. Specifically, this study provides valuable insight that female service members are a population of concern for the female athlete triad, especially among female veterans.

## Data Availability

All data produced in the present study are available upon reasonable request to the authors

## Appendix

**Figure 1:**
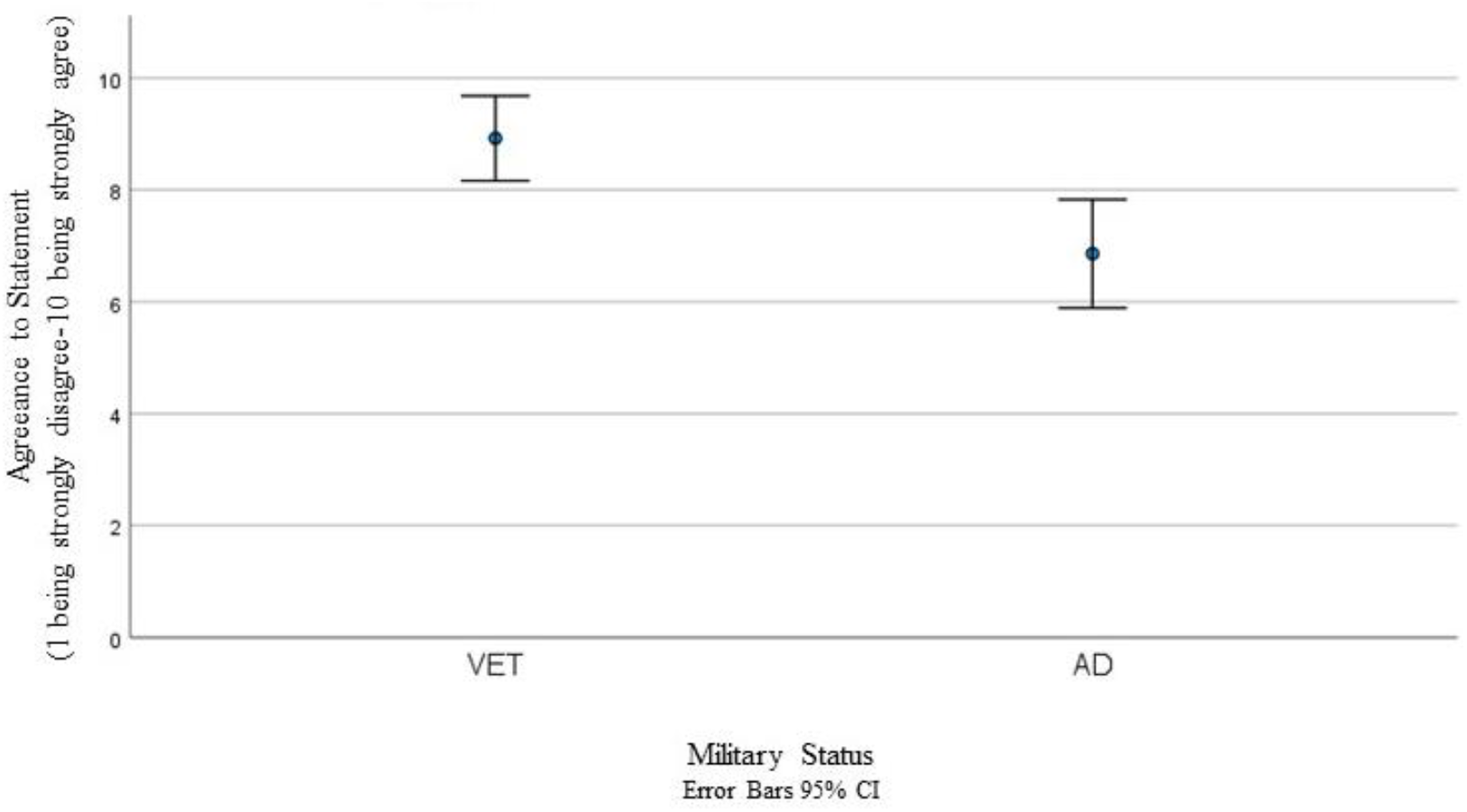
Difference between Female Veterans and Active Duty Service Members in Response to ““I Believe Most of My Eating Habits Have Been Influenced by the Diet Culture in the Army” (Q1)

**Figure 2:**
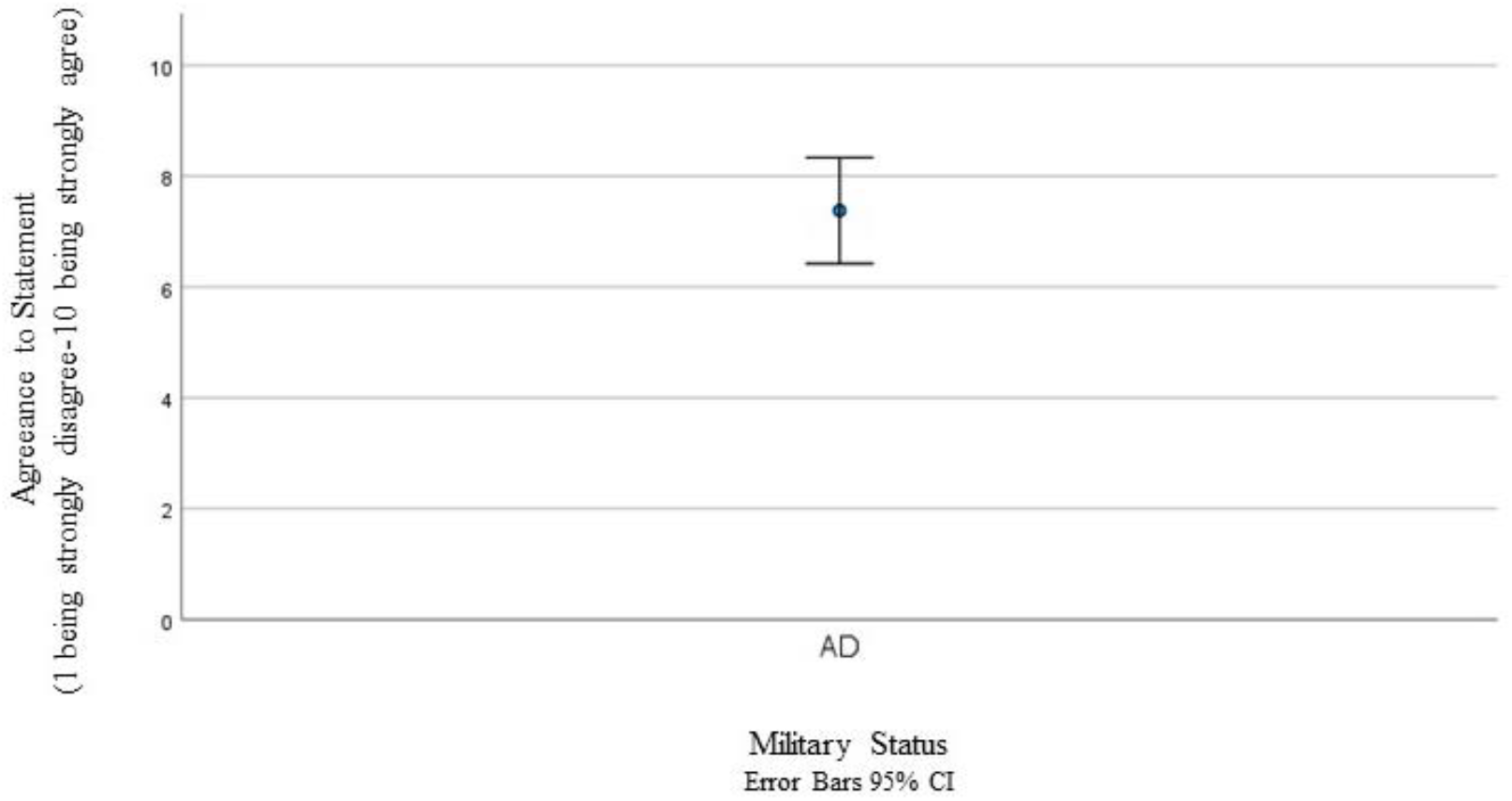
Active Duty Service Members’ Response To ”“I am concerned about meeting Army height and weight requirements” (Q2)

**Figure 3:**
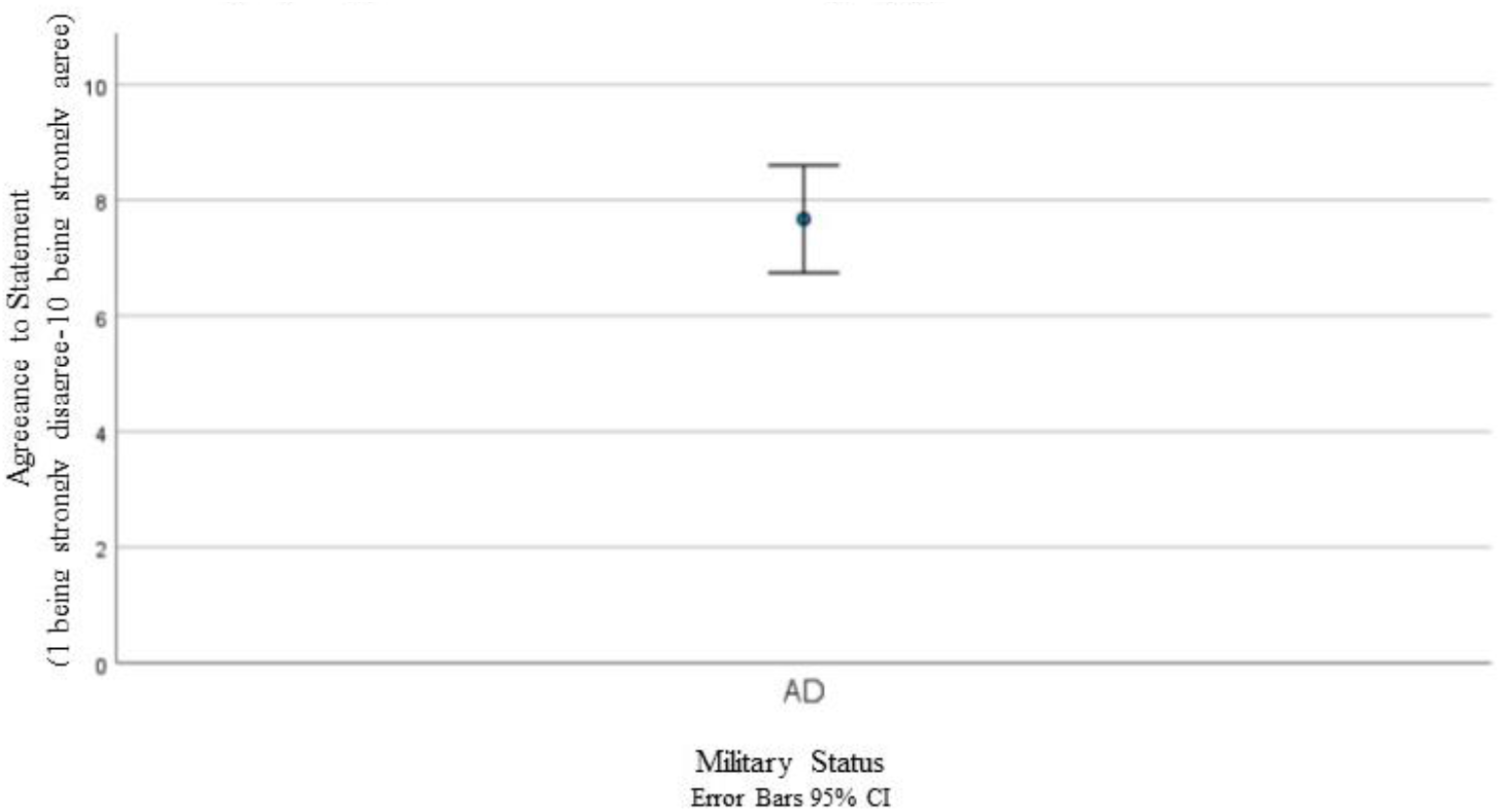
Responses from Female Active- Duty Service Members to “I Have Done Things to Keep My weight Down that I Believe are Unhealthy” (Q3)

## Footnotes

1 Alexiaa Sim and Stephen F. Burns, “Review: Questionnaires as Measures for Low Energy Availability (LEA) and Relative Energy Deficiency in Sport (RED-S) in Athletes,” *Journal of Eating Disorders* 9, no. 1 (March 31, 2021): https://doi.org/10.1186/s40337-021-00396-7.

2 Thomas J. O’leary et al., “Sex Differences in Neuromuscular Fatigability in Response to Load Carriage in the Field in British Army Recruits,” *Journal of Science and Medicine in Sport* 21, no. 6 (June 2018): [Page #], https://doi.org/10.1016/j.jsams.2017.10.018.

3 Nattiv Aurelia et al., “The Female Athlete Triad,” *Medicine & Science in Sports & Exercise* 39, no. 10 (October 2007): [Page #], https://doi.org/10.1249/mss.0b013e318149f111.

4 Gaston P. Bathalon et al., “The Effect of Proposed Improvements to the Army Weight Control Program on Female Soldiers,” *Military Medicine* 171, no. 8 (2006).

5 Melin A, Tornberg AB, Skouby S, et al. The LEAF questionnaire: a screening tool for the identification of female athletes at risk for the female athlete triad. Br J Sports Med. 2014;48(7):540–5.

6 McNulty KY, Adams CH, Anderson JM, et al. Development and validation of a screening tool to identify eating disorders in female athletes. J Am Diet Assoc. 2001;101(8):886–92.

7 Kimberly A. Arditte hall et al., “Eating Disorder Symptoms in Female Veterans: The Role of Childhood, Adult, and Military Trauma Exposure.,” *Psychological Trauma: Theory, Research, Practice, and Policy* 10, no. 3 (May 2018): [Page #], https://doi.org/10.1037/tra0000301.

